# NEXIM: A Nash Equilibrium-Based Framework for Stable Explainable AI in Medical Applications

**DOI:** 10.64898/2026.06.25.26356568

**Authors:** Dipak P. Upadhyaya, Deepak K. Gupta, Katrina Prantzalos, Pedram Golnari, Vivikta Lyer, Cole Zweber, Subhashini Sivagnanam, Amitava Majumdar, Satya S. Sahoo

## Abstract

Reliable explanations are important for trustworthy medical applications of artificial intelligence (AI), but attribution-based explanations can vary across model randomization and small analytic changes. We present NEXIM (Nash Equilibrium-based Explainability and Interpretability Model), implemented here as an accuracy-constrained, equilibrium-inspired model-selection framework that jointly evaluates held-out prediction error, explanation stability, and cross-model connectivity. The implementation evaluated ten GradientBoostingRegressor models per prediction horizon, differing only by random seed (0-9), using a fixed 75/25 patient split. Kernel SHAP attribution vectors were compared using Spearman rank correlation, and graph connectivity summarized whether each model belonged to a dense explanation-similarity region. Candidate models within 0.02 Montreal Cognitive Assessment points of the best root mean squared error (RMSE) were ranked using a multiplicative Explanation Equilibrium Score. In longitudinal Parkinson’s Progression Markers Initiative data, NEXIM selected the RMSE-optimal model at the one- and three-year horizons. At the two-year horizon, it selected Model 4 rather than the RMSE-only Model 8, increasing scaled stability from 0.8757 to 0.8847 and normalized graph connectivity from 0.889 to 1.000 while increasing RMSE by only 0.0014. The two models retained the same top-20 feature set but differed modestly in feature order, illustrating that NEXIM primarily acted as a reproducibility screen rather than identifying clinically contradictory explanations. Stability and consensus are treated as reproducibility criteria, not evidence of causal faithfulness, clinical usefulness, or improved patient outcomes. NEXIM may therefore serve as a governance checkpoint for model refresh and documentation, but external validation, stronger model-family baselines, and prospective clinical evaluation remain necessary.

## 1. Introduction

Artificial intelligence (AI) is increasingly used to support clinical prediction, disease-risk assessment, and longitudinal monitoring of patient outcomes. As these systems become integrated into healthcare environments, their value depends not only on predictive accuracy but also on whether model behavior can be documented, reproduced, and interpreted appropriately. Human-centered research in clinical explainable artificial intelligence (XAI) emphasizes that technical explanation metrics should be complemented by evaluation of clinician understanding, trust calibration, usability, and task-specific clinical relevance.^1-5^

Most machine-learning pipelines still select models primarily by predictive performance and evaluate interpretability only after model training. This creates a reliability problem: models with nearly identical predictive error may produce different feature rankings or attribution patterns across random seeds, model configurations, or small analytic perturbations.^6,7^ Such explanation instability is especially important in clinical settings, where explanations are often reviewed as ranked predictors or influential clinical features rather than as full attribution vectors. If similarly accurate models emphasize different predictors, the resulting explanations may be difficult to reproduce, document, or compare during model review.

Attribution-based explanation methods, including SHapley Additive exPlanations (SHAP) and local surrogate approaches such as LIME, provide useful summaries of model behavior and are widely used in clinical machine learning.^8-10^ However, explanation consistency should not be equated with explanation quality. Rank stability and feature-overlap metrics assess reproducibility, whereas faithfulness to the fitted model, causal validity, clinical actionability, clinician usability, and impact on decision making require separate evaluation.^2,4,5,11,12,13^ In this study, we therefore use the term explanation reliability narrowly to describe reproducibility and agreement of attribution patterns across candidate models.

To address this limitation, we introduce NEXIM (Nash Equilibrium-based Explainability and Interpretability Model) implemented here as an accuracy-constrained, equilibrium-inspired model-selection framework. NEXIM jointly evaluates held-out prediction error, explanation stability, and cross-model explanation connectivity. Predictive performance is treated as the primary constraint, and explanation reproducibility is used to distinguish models whose RMSE values are practically equivalent. The implemented framework is not a formal Nash-equilibrium solver; rather, it uses a transparent multiplicative Explanation Equilibrium Score (EES) to identify candidate models that balance predictive adequacy with reproducible global attribution profiles.

We evaluated NEXIM using longitudinal Parkinson’s Progression Markers Initiative (PPMI) data to predict Montreal Cognitive Assessment (MoCA) outcomes at three follow-up horizons: MCATOT_V04, MCATOT_V06, and MCATOT_V08. Parkinson’s disease provides an informative clinical setting because cognitive decline is longitudinal, heterogeneous, and commonly modeled using demographic, clinical, cognitive, imaging, behavioral, psychiatric, and genetic variables. The analysis asks a focused question: when random-seed variants of the same model family have similar held-out RMSE, can an accuracy-constrained explanation-reliability score identify a model whose SHAP attribution profile is more representative of the candidate ensemble?

This study makes three contributions. First, it presents a reproducible workflow for reporting predictive accuracy and explanation stability together during model selection. Second, it defines an explicit Explanation Equilibrium Score with documented RMSE and graph-connectivity thresholds rather than undocumented weighting choices. Third, it provides a longitudinal PPMI case study demonstrating both the utility and the limits of stability-aware model selection as an exploratory governance tool.

## 2. Methods

### 2.1 Overview of NEXIM Framework

NEXIM is a framework designed to identify predictive models whose explanations are reliable, stable, and reproducible across candidate configurations. The central idea is to evaluate predictive performance and explanation reproducibility jointly while preserving predictive adequacy as the primary constraint. NEXIM does not assume that stable explanations are causally correct or clinically useful; instead, it treats stability and consensus as reproducibility criteria that should be evaluated before a model is selected for clinical interpretation. The framework consists of four stages: training multiple candidate predictive models, generating feature-attribution explanations, computing explanation stability and connectivity metrics, and selecting an eligible near-tie model using an accuracy-constrained Explanation Equilibrium Score.^14^

**Figure 1.**
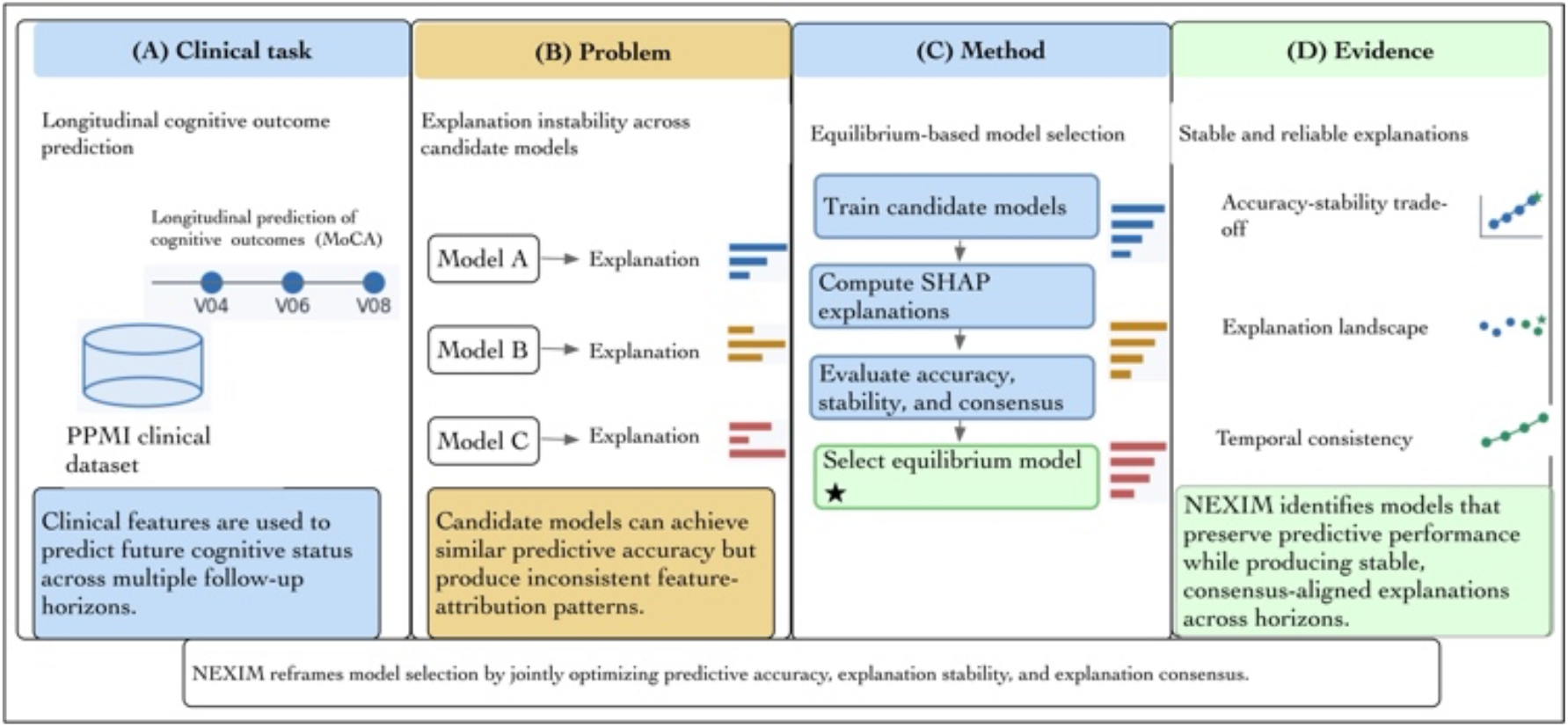
Conceptual overview of the NEXIM framework. NEXIM addresses explanation instability by selecting models that meet predictive adequacy criteria while improving explanation reproducibility and cross-model explanation connectivity. Candidate models are evaluated using held-out RMSE, SHAP rank stability, and graph connectivity, and eligible near-tie models are ranked using the multiplicative Explanation Equilibrium Score.

### 2.2 Predictive Modeling

For each horizon, all 301 participants had an observed outcome. Data were split once into 225 training participants and 76 held-out participants using train_test_split(test_size=0.25, random_state=42). Missing predictor values were median-imputed after categorical variables were one-hot encoded. The prespecified feature list contained 214 baseline variables and expanded to 229 model inputs after encoding. Ten scikit-learn GradientBoostingRegressor models were trained per horizon with identical default hyperparameters and random_state values 0 through 9. Thus, the candidate set tests random-seed variability within one model family. Predictive performance was measured by held-out RMSE in MoCA points, consistent with continuous-outcome clinical prediction evaluation.^15^

### 2.3 Explanation Generation

Kernel SHAP was used to explain each fitted regressor using Shapley-value-based feature attribution.^9,16^ Because the held-out set contained 76 participants, all 76 were used as the background sample and explanation sample, with nsamples=200. For each model, absolute SHAP values were averaged across held-out participants and normalized to sum to one, producing a global attribution vector over all the encoded inputs. These global vectors, not patient-level explanations, were used for model-to-model comparisons.

### 2.4 Explanation Stability

Explanation stability quantifies the extent to which different candidate models produce consistent attribution patterns. For each pair of candidate models, stability was computed as the Spearman rank correlation between their normalized SHAP attribution vectors. Spearman correlation was selected because it evaluates agreement in feature ranking rather than absolute attribution magnitude, which is appropriate when clinicians review explanations as ordered lists of influential predictors.^16^ For each model, the overall stability score was computed as the mean correlation between its attribution vector and those of all other models in the candidate ensemble. Higher stability therefore indicates that a model’s explanation aligns with the broader ensemble rather than representing an isolated or idiosyncratic attribution pattern.

### 2.5 Model Consensus

Two complementary consensus summaries were used. First, descriptive top-feature overlap was calculated with Jaccard similarity for the top 20 features. Second, the primary selection pipeline constructed an undirected graph using scaled pairwise similarity (rho+1)/2; an edge was added when similarity was at least 0.85. Normalized degree centrality, rescaled by the maximum degree within a horizon, defined the topology score T_topo. T_topo therefore measures ensemble connectivity and should not be interpreted as pairwise Jaccard overlap.^5,17^

### 2.6 Equilibrium-Based Model Selection

Model selection in NEXIM was implemented as an accuracy-constrained multi-criteria rule. Within each horizon, predictive optimality was min-max scaled as P_pred = 1 - (RMSE - RMSE_min)/(RMSE_max - RMSE_min). Models were eligible only when RMSE was no more than 0.02 MoCA points above the best observed RMSE. This tolerance made predictive performance the primary constraint while allowing explanation reliability to break near-ties. For eligible models, the multiplicative EES combined P_pred, S_stab, and T_topo as described below. The RMSE tolerance and graph-similarity threshold are tunable quantities that should be assessed through sensitivity analyses before deployment.^14^

### 2.7 Theoretical Formulation of NEXIM

For an eligible model m, the implemented Explanation Equilibrium Score was

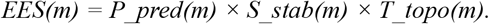

The eligible model with the largest EES was selected. The multiplicative form has no 0.50/0.30/0.20 weights: a low value in any component reduces the final score, and the RMSE tolerance prevents a reliability gain from compensating for a clinically meaningful loss of accuracy. The tunable quantities are the RMSE tolerance (0.02 in this analysis) and graph-similarity threshold (0.85). Sensitivity to these values should be evaluated before deployment.

This operational definition is equilibrium-inspired rather than a formal game-theoretic Nash equilibrium.^18-20^ No strategic players, payoff functions, or best-response dynamics were estimated. We retain the NEXIM name for continuity while limiting the claim to balanced, accuracy-constrained model selection. The selection procedure was applied independently at each prediction horizon; the final choice was based on RMSE eligibility and maximum multiplicative EES.

### 2.8 Experimental Evaluation

The framework was evaluated on de-identified longitudinal PPMI data to predict MoCA at MCATOT_V04, MCATOT_V06, and MCATOT_V08. These horizons provide a case study of whether explanation rankings remain reproducible as the interval between baseline predictors and outcomes increases. Primary comparisons included RMSE-only selection, direct highest-stability selection, and NEXIM selection. We report exact changes in RMSE, S_stab, T_topo, and feature ranks. The available analysis used one held-out split and did not produce a defensible patient-level inferential comparison of selection rules; therefore, stability differences are interpreted descriptively rather than as statistically significant effects.

### 2.9 Implementation and Statistical Analysis

Analyses were implemented in Python using Pandas, NumPy, scikit-learn, SHAP, and NetworkX. Random seeds, split parameters, Kernel SHAP sample count, RMSE tolerance, and graph threshold are reported above to support reproduction. The current evidence is configuration-level and exploratory; external validation and repeated nested resampling are needed for formal uncertainty estimates.^21^

### 2.10 Data Governance and Ethics

This study used de-identified PPMI data under the applicable PPMI Data Use Agreement.^22, 23^ Analyses were limited to the approved research purpose and followed PPMI restrictions against participant re-identification, unauthorized redistribution, and use of participant-level data in tools or computing environments that could retain, disclose, or train on the data without appropriate containment guarantees. No attempt was made to contact participants or recover protected identifiers. All results are reported in aggregate form. Because the analysis used secondary de-identified data, the study did not involve direct participant contact.

## 3. Results

### 3.1 Cohort Characteristics and Prediction Tasks

The analytic cohort consisted of 301 participants from the PPMI with longitudinal clinical assessments and cognitive outcomes measured using the MoCA. After preprocessing and feature harmonization, baseline demographic, clinical, cognitive, imaging, behavioral, psychiatric, and genetic variables were used to construct predictive models of future cognitive performance. The dataset was randomly divided into training and testing subsets using a 75/25 split to ensure that model development and evaluation were performed on independent samples. Prediction tasks were defined for three longitudinal horizons corresponding to MCATOT_V04, MCATOT_V06, and MCATOT_V08, representing cognitive outcomes approximately one, two, and three years after baseline evaluation. Table 1 summarizes the dataset characteristics, prediction targets, and explainability components used within the NEXIM framework.

**Table 1.** Dataset characteristics and variables used in the NEXIM framework.

| Component | Description |
| --- | --- |
| Data source | Parkinson's Progression Markers Initiative |
| Cohort | 301 participants for each prediction horizon |
| Split | 225 training / 76 held-out participants (75/25; random_state=42) |
| Candidate models | 10 GradientBoostingRegressor models; seeds 0-9; identical default hyperparameters |
| Feature domains | Demographic, clinical, cognitive, imaging, behavioral, psychiatric, and genetic variables |
| Representative features | Age, disease duration, MCATOT_V00, DATSCAN measures, ESS_total, GDS_total, APOE ε4 |
| Prediction targets | MCATOT_V04, MCATOT_V06, MCATOT_V08 |
| Explanation method | Kernel SHAP |
| Stability metric | Scaled mean pairwise Spearman correlation, $S_{stab} = (\text{mean } \rho + 1)/2$ |
| Consensus/connectivity metric | Graph degree centrality at scaled similarity $\geq 0.85$; top-20 Jaccard reported descriptively |
| Selection criterion | Multiplicative Explanation Equilibrium Score |
| Model selection objective | Preserve predictive adequacy while favoring reproducible explanation structure |

### 3.2 Accuracy-Stability Trade-off and NEXIM Equilibrium Selection

Table 2 compares RMSE-only selection, direct highest-stability selection, and NEXIM. At MCATOT_V04, Model 5 was both the RMSE-only and NEXIM choice. At MCATOT_V08, Model 1 had the best RMSE and the highest stability and was therefore selected by all three rules. The only divergence occurred at MCATOT_V06: NEXIM selected Model 4 instead of RMSE-only Model 8. RMSE increased from 2.1414 to 2.1428 (+0.0014 MoCA points; approximately 0.06%), while S_stab increased from 0.8757 to 0.8847 (+0.0090; approximately 1.03%) and T_topo increased from 0.889 to 1.000. These are modest descriptive differences, not evidence of a significant stability improvement across all horizons.

**Table 2.** Model Performance and Equilibrium Selection.

| Horizon | Selection rule | Model | RMSE | S_stab | T_topo | EES |
| --- | --- | --- | --- | --- | --- | --- |
| V04 | Best RMSE | 5 | 2.2997 | 0.8962 | 1.00 | 0.8962 |
|  | Highest Stability | 3 | 2.3138 | 0.9018 | 1.00 | 0.6208 |
|  | NEXIM | 5 | 2.2997 | 0.8962 | 1.00 | 0.8962 |
| V06 | Best RMSE | 8 | 2.1414 | 0.8757 | 0.89 | 0.7784 |
|  | Highest Stability | 3 | 2.1851 | 0.8946 | 1.00 | 0.4897 |
|  | NEXIM | 4 | 2.1428 | 0.8847 | 1.00 | 0.8721 |
| V08 | Best RMSE | 1 | 2.1467 | 0.8856 | 1.00 | 0.8856 |
|  | Highest Stability | 1 | 2.1467 | 0.8856 | 1.00 | 0.8856 |
|  | NEXIM | 1 | 2.1467 | 0.8856 | 1.00 | 0.8856 |

Across selected models, age and baseline MoCA were consistently prominent. Disease duration remained within the top 20 but changed rank across horizons, and biomarker or clinical variables such as neurofilament light, phosphorylated tau, autonomic measures, and sleep-related features showed greater rank variation. This pattern supports clinical plausibility assessment while underscoring that attribution stability alone does not establish biological validity.^24-26^

**Figure 2.**
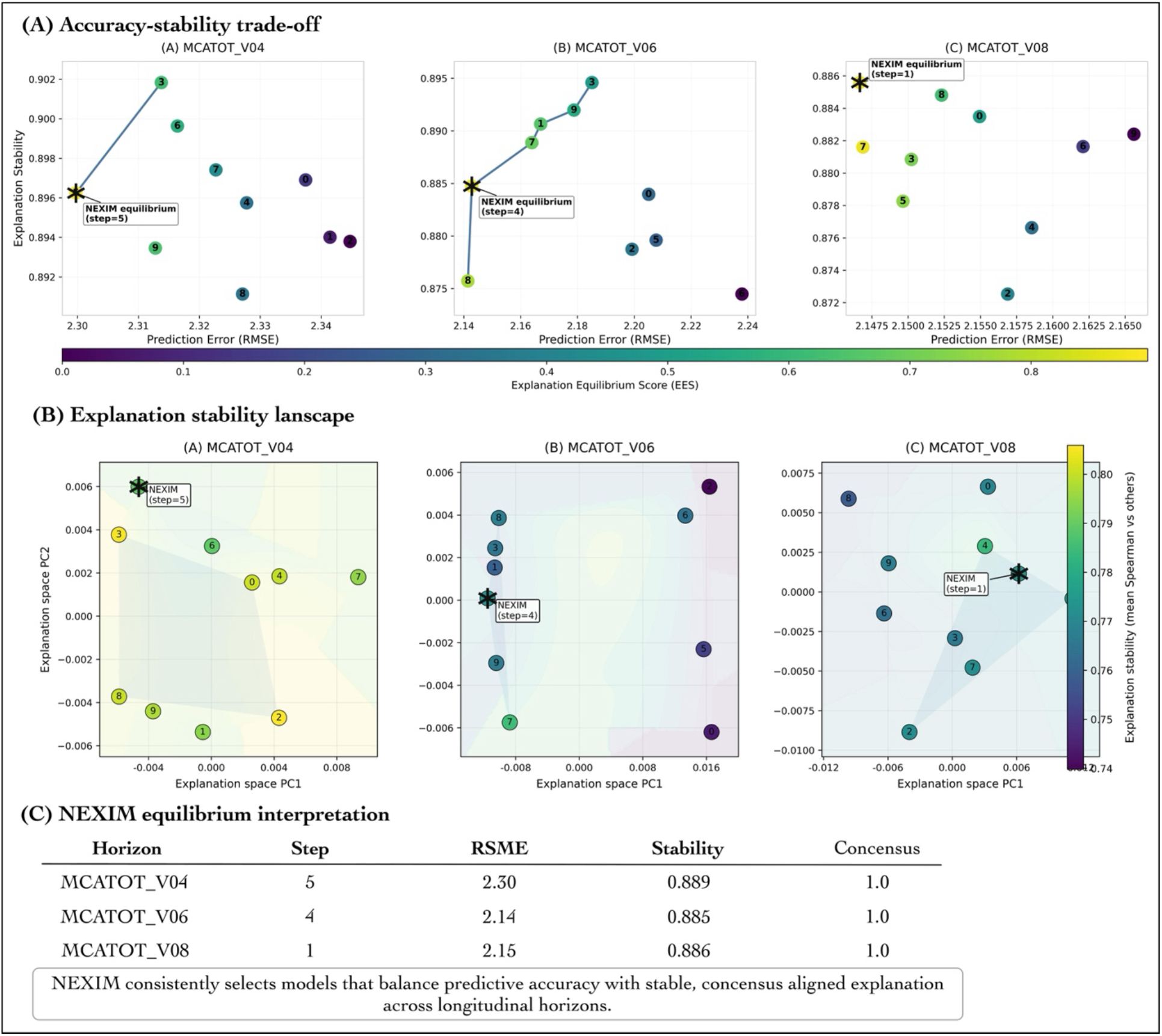
Accuracy-stability trade-off across candidate random-seed configurations.

Each panel shows held-out RMSE and scaled explanation stability for ten GradientBoostingRegressor seeds. Asterisk markers denote the NEXIM-selected model among candidates within 0.02 RMSE points of the best model. The plotted lines and point locations illustrate trade-offs; selection used the documented RMSE-eligibility rule and multiplicative Explanation Equilibrium Score.

### 3.3 Temporal Stability of Feature Attributions

Figure 3 presents normalized SHAP attribution values for the most influential predictors across the MCATOT_V04, MCATOT_V06, and MCATOT_V08 prediction tasks. Several predictors demonstrated stable explanatory influence across horizons. Age, baseline cognitive performance (MCATOT_V00), and disease duration consistently ranked among the most influential predictors across all prediction tasks. Although attribution magnitudes varied moderately across prediction horizons, the relative ranking of key predictors remained largely consistent. Age exhibited increasing attribution influence at longer prediction horizons, reflecting the progressive role of aging in cognitive decline. Baseline cognitive performance contributed strongly across intermediate horizons, while certain clinical biomarkers showed greater influence in earlier prediction stages.

**Figure 3.**
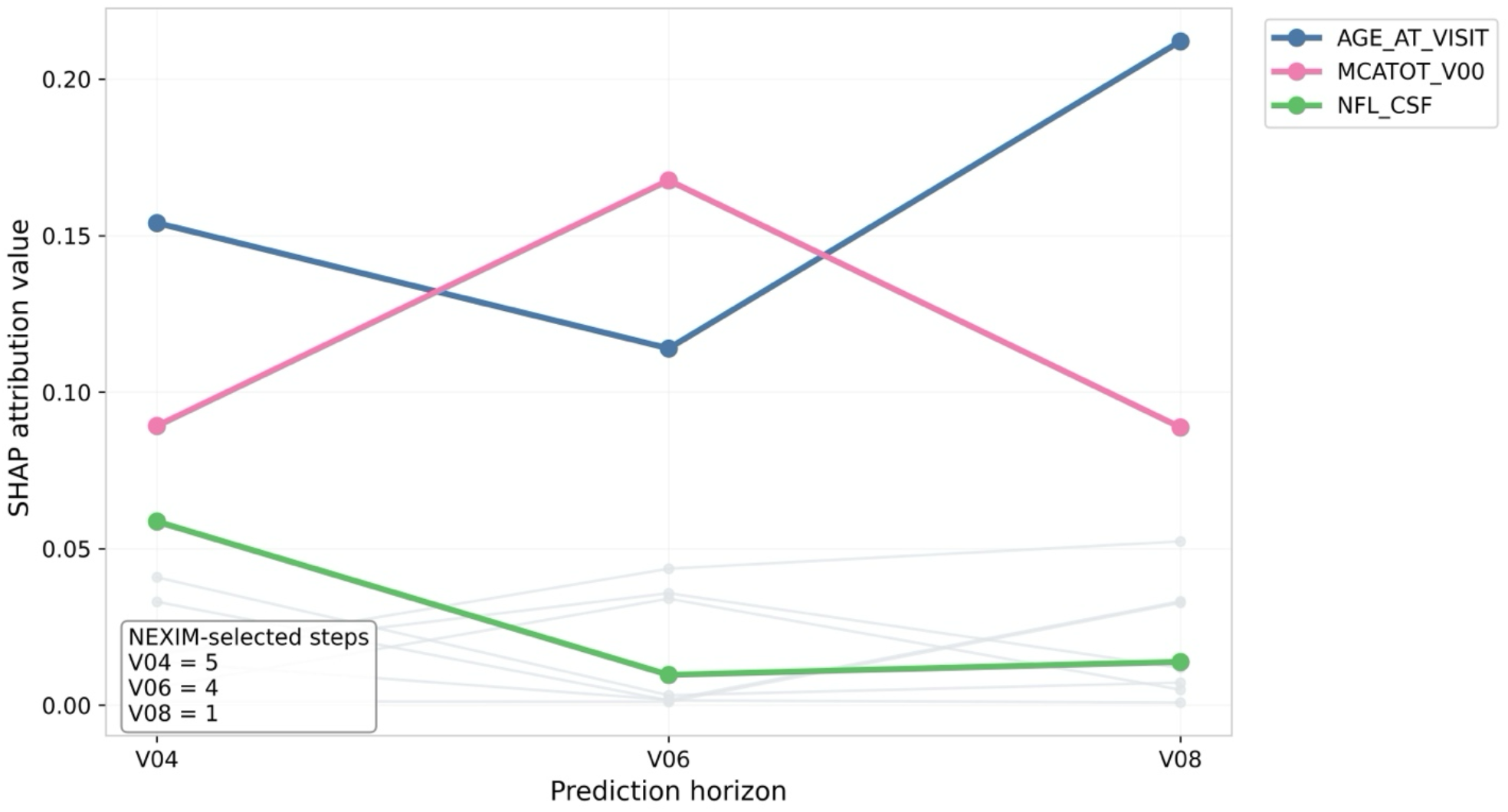
Temporal trajectories of normalized SHAP attribution values for key predictors across prediction horizons. Colored lines highlight selected high-attribution predictors, while thin gray lines represent additional lower-ranked predictors included to show background attribution variability across horizons. The inset reports the NEXIM-selected model at each prediction horizon.

### 3.4 Ensemble Connectivity and a Concrete Rank-Order Example

At MCATOT_V06, the RMSE-only Model 8 and NEXIM Model 4 provide the most informative near-tie. Their saved top-20 feature sets were identical (pairwise Jaccard=1.00), so they did not present contradictory biomarker sets. They did, however, reorder several features: Model 4 ranked phosphorylated tau above temperature and moved sleep-limb-movement, dream-verbalization, and rhinoceros-naming variables upward, whereas Model 8 ranked constipation-related symptoms higher. Table 3 reports the top ten. This example clarifies that the observed disagreement was modest and rank-based. The difference in T_topo (1.000 for Model 4 versus 0.889 for Model 8) reflects connectivity to the full ten-model similarity graph, not disagreement between these two models’ top-20 sets. Model 4 occupied a maximally connected region of the candidate ensemble, while Model 8 lacked one qualifying graph connection.

**Table 3.**
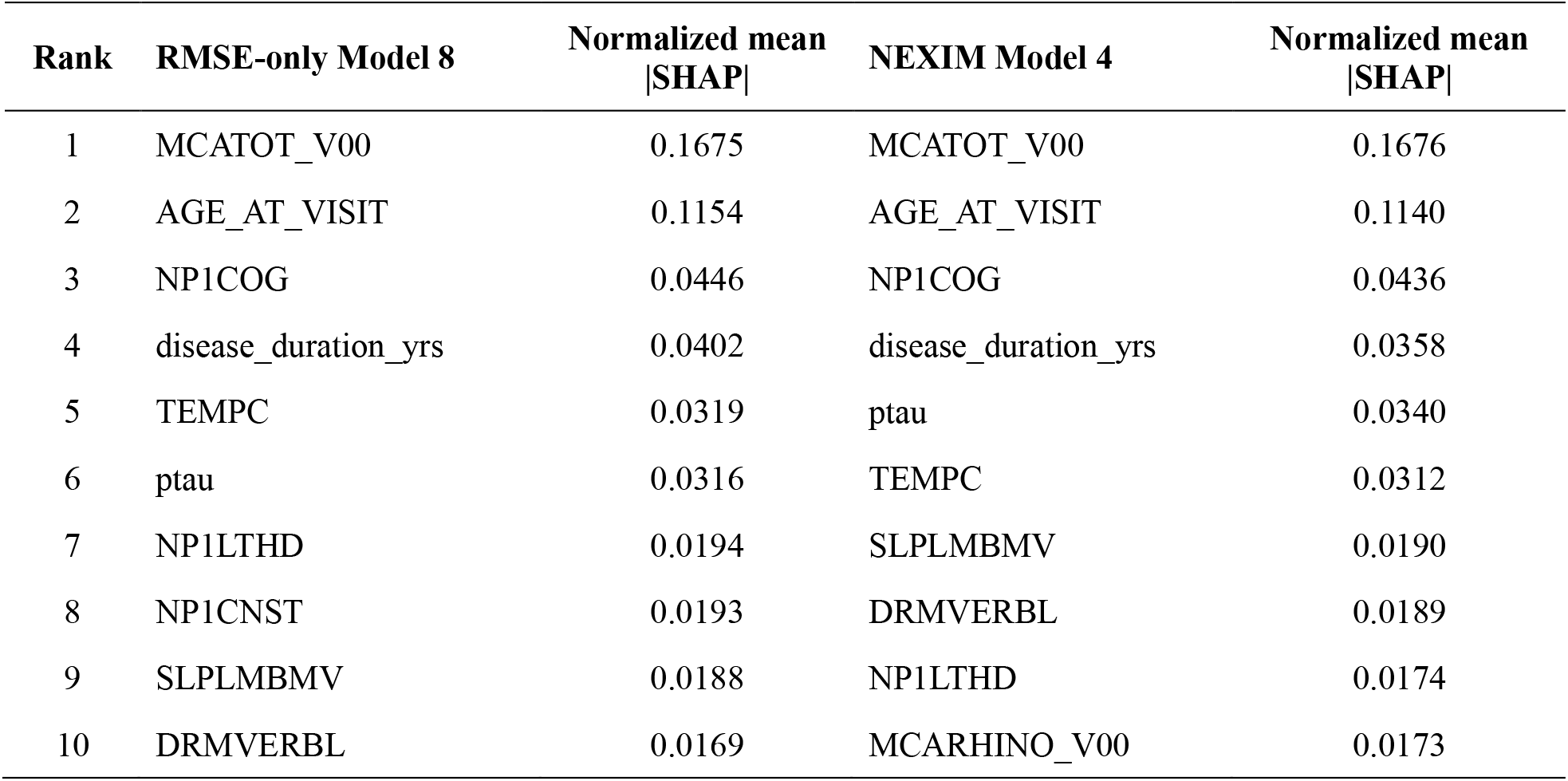
Top-ten attribution rankings for the two near-equivalent MCATOT_V06 models.

### 3.5 Explanation Disagreement Across Candidate Models

Figure 4 visualizes explanation-similarity graphs for prediction horizons. In these networks, nodes represent candidate models and edges represent similarity between SHAP attribution vectors. Candidate models formed distinct communities corresponding to groups of models with similar explanation structures. Importantly, NEXIM-selected models were consistently located within densely connected communities characterized by strong intra-community agreement. This pattern suggests that equilibrium models correspond to stable explanation regimes rather than isolated attribution patterns.

**Figure 4.**
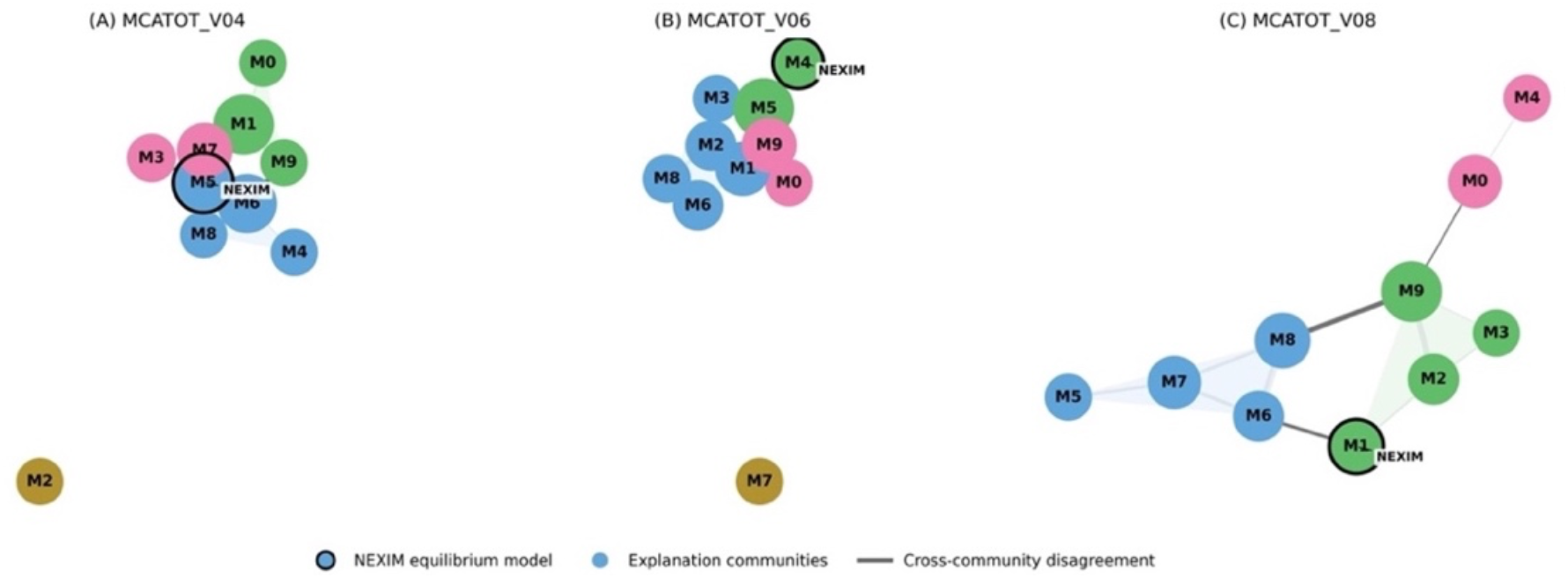
Model explanation disagreement networks across prediction horizons. Nodes represent candidate models and edges represent similarity between SHAP attribution vectors. Colors denote explanation communities identified through network clustering. NEXIM-selected equilibrium models appear within dense communities, indicating stable explanation regimes.

### 3.6 Feature-Level Stability Analysis

Figure 5 presents a heatmap of feature attribution stability across candidate models and prediction horizons. Several predictors exhibited consistently stable attribution patterns across models, including age, baseline cognitive score, and disease duration. These predictors represent clinically plausible determinants of cognitive decline and therefore provide interpretable explanatory signals. In contrast, certain biomarker variables showed greater variability in attribution magnitude across candidate models, indicating sensitivity to model configuration. These results highlight the importance of evaluating explanation stability when interpreting feature importance in clinical machine-learning models.

**Figure 5.**
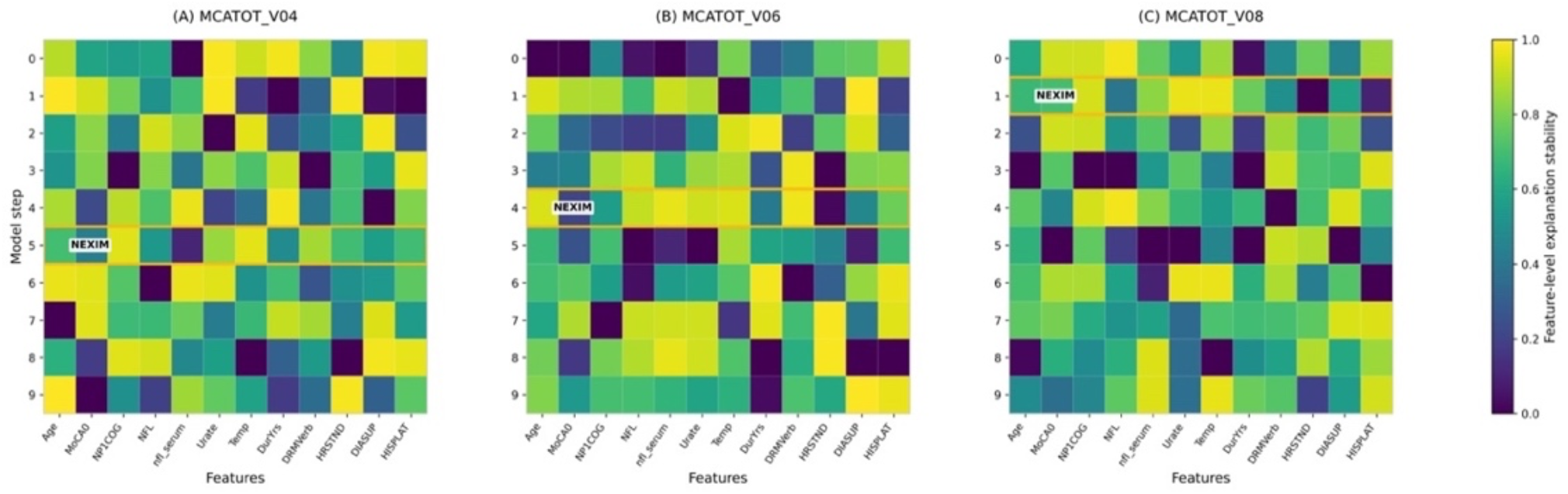
Explanation stability heatmap across prediction horizons. Rows correspond to candidate models and columns represent explanatory features ranked by attribution importance. Cell values indicate cross-model stability of feature attributions. Highlighted rows indicate NEXIM-selected equilibrium models.

### 3.7 Statistical Interpretation and Sensitivity Limitations

The present study used one held-out split and ten random-seed variants of a single model family. Consequently, the analysis supports descriptive comparison of RMSE, S_stab, T_topo, and feature-rank patterns, but not formal patient-level inference about superiority of one selection rule. Available perturbation and bootstrap artifacts were not sufficient to support a comparative confidence interval or p-value across selection rules. We therefore report the MCATOT_V06 difference as a descriptive near-tie and interpret NEXIM as an exploratory reproducibility screen. Future confirmatory analyses should use repeated or nested resampling, patient-level bootstrap differences, sensitivity analyses for the 0.02 RMSE tolerance and 0.85 graph threshold, and external validation cohorts.

## Discussion

This study introduced NEXIM, an equilibrium-based framework that integrates explanation reliability directly into model selection. The main finding is modest but useful: among ten random-seed variants of the same GradientBoostingRegressor family, NEXIM preserved the RMSE-optimal choice at the one-and three-year MoCA horizons and selected a near-equivalent, more ensemble-connected model at the two-year horizon. At MCATOT_V06, Model 4 increased RMSE by only 0.0014 MoCA points relative to RMSE-only Model 8 while improving scaled explanation stability from 0.8757 to 0.8847 and graph connectivity from 0.889 to 1.000. Thus, when predictive performance is tied, explanation reproducibility can serve as a secondary criterion for selecting a model whose attribution profile is more representative of the candidate ensemble.

These findings should be interpreted as improved reproducibility, not proof of improved explanation correctness, causal faithfulness, clinical utility, or patient benefit. Prior studies show that feature-attribution explanations can vary across model configurations and analytic perturbations despite similar predictive performance.^6,7^ NEXIM addresses this disagreement problem by comparing global SHAP attribution rankings across candidate models, but rank stability and top-feature overlap are only reproducibility measures. They do not establish faithfulness to the fitted model, causal validity, clinical actionability, or effects on clinician reliance.^2,4,5,11,12^ This distinction directly motivates future evaluations that combine stability metrics with faithfulness tests, expert review, and clinician-facing usability studies. The MCATOT_V06 near-tie also clarifies the clinical interpretation of NEXIM. Model 4 and Model 8 shared the same top-20 features, so the observed disagreement was not a contradictory biomarker signal. Instead, differences were rank-based: Model 4 ranked phosphorylated tau above temperature and moved sleep-limb-movement, dream-verbalization, and rhinoceros-naming features upward, whereas Model 8 ranked constipation-related symptoms higher. Age and baseline MoCA remained dominant across horizons, and disease duration stayed within the top-20 features despite rank changes. These patterns are consistent with Parkinson’s disease cognition literature linking demographic, baseline cognitive, clinical, and multimodal factors to cognitive impairment and longitudinal decline.^24,25^ However, clinical plausibility is not validation; stable predictors may still reflect cohort structure, confounding, measurement effects, or model-specific behavior.

NEXIM may be most useful as a Learning Health System and MLOps governance checkpoint. During model refresh, deployment teams could require acceptable predictive performance, then flag material explanation changes for documentation and review.^12,27^ The current study also defines important limits. The candidates differed only by random seed within one model family, the analysis used a single 75/25 split with 76 held-out participants, and the available bootstrap artifacts did not support patient-level inferential comparisons. Kernel SHAP global attribution vectors summarize average feature influence but do not measure patient-level usefulness or causal faithfulness.^8,15^ Future work should compare heterogeneous model families, test RMSE-tolerance and graph-threshold sensitivity, use repeated or nested resampling with bootstrap differences,^21^ validate externally, and evaluate whether stability-aware selection improves clinician understanding, trust calibration, utilization, or outcomes.

## Conclusion

NEXIM provides a transparent framework for using explanation reproducibility as a secondary model-selection criterion after predictive adequacy is established. In this PPMI case study, NEXIM preserved the RMSE-optimal model at the one- and three-year MoCA prediction horizons and selected a more ensemble-connected near-tie at the two-year horizon with negligible loss in predictive performance. These findings support NEXIM as an exploratory governance and documentation tool for stability-aware clinical AI development. However, explanation stability should not be interpreted as evidence of causal validity, explanation faithfulness, clinical usefulness, or improved patient outcomes. Future work should evaluate NEXIM across heterogeneous model families, repeated resampling designs, external cohorts, alternative explanation methods, and prospective clinical AI workflows before deployment.

## Data Availability

All data produced in the present study are available upon reasonable request to the authors

https://www.ppmi-info.org/

## References

1. Wiens J, Saria S, Sendak M, Ghassemi M, Liu VX, Doshi-Velez F, et al. Do no harm: a roadmap for responsible machine learning for health care. Nature medicine. 2019;25(9):1337–40.

2. Amann J, Blasimme A, Vayena E, Frey D, Madai VI, Consortium PQ. Explainability for artificial intelligence in healthcare: a multidisciplinary perspective. BMC medical informatics and decision making. 2020;20(1):310.

3. Tonekaboni S, Joshi S, McCradden MD, Goldenberg A, editors. What clinicians want: contextualizing explainable machine learning for clinical end use. Machine learning for healthcare conference; 2019: PMLR.

4. Nicolson A, Bradburn E, Gal Y, Papageorghiou AT, Noble JA. The human factor in explainable artificial intelligence: clinician variability in trust, reliance, and performance. npj Digital Medicine. 2025;8(1):658.

5. Pahud de Mortanges A, Luo H, Shu SZ, Kamath A, Suter Y, Shelan M, et al. Orchestrating explainable artificial intelligence for multimodal and longitudinal data in medical imaging. NPJ digital medicine. 2024;7(1):195.

6. Haibe-Kains B, Adam GA, Hosny A, Khodakarami F, Massive Analysis Quality Control (MAQC) Society Board of Directors, Waldron L, et al. Transparency and reproducibility in artificial intelligence. Nature. 2020;586(7829):E14–E16. doi:10.1038/s41586-020-2766-y.

7. Bhatt U, Weller A, Moura JM. Evaluating and aggregating feature-based model explanations. arXiv preprint arXiv:2005.00631. 2020.

8. Krishna S, Han T, Gu A, Wu S, Jabbari S, Lakkaraju H. The disagreement problem in explainable machine learning: A practitioner’s perspective. arXiv preprint arXiv:2202.01602. 2022.

9. Lundberg SM, Lee SI. A Unified Approach to Interpreting Model Predictions. Advances in Neural Information Processing Systems. 2017;30.

10. Ribeiro MT, Singh S, Guestrin C. “Why Should I Trust You?” Explaining the Predictions of Any Classifier. Kdd’16: Proceedings of the 22nd Acm Sigkdd International Conference on Knowledge Discovery and Data Mining. 2016:1135–44.

11. Rajkomar A, Dean J, Kohane I. Machine Learning in Medicine. Reply. N Engl J Med. 2019;380(26):2589–90.

12. Rudin C. Stop explaining black box machine learning models for high stakes decisions and use interpretable models instead. Nat Mach Intell. 2019;1(5):206–15.

13. Rudin C, Chen CF, Chen Z, Huang HY, Semenova L, Zhong CD. Interpretable machine learning: Fundamental principles and 10 grand challenges. Stat Surv. 2022;16:1–85.

14. Deb K. Multi-objective optimisation using evolutionary algorithms: an introduction. Multi-objective evolutionary optimisation for product design and manufacturing: Springer; 2011. p. 3–34.

15. Steyerberg EW. Clinical prediction models: Springer; 2019.

16. Molnar C. Interpretable Machine Learning: A Guide For Making Black Box Models Explainable. 2nd ed. 2022.

17. Bhatt U, Xiang A, Sharma S, Weller A, Taly A, Jia Y, et al., editors. Explainable machine learning in deployment. Proceedings of the 2020 conference on fairness, accountability, and transparency; 2020.

18. Watson J. Strategy: An Introduction to Game Theory. 3rd ed. New York: W. W. Norton & Company; 2013.

19. Leyton-Brown K, Shoham Y. Essentials of game theory: A concise multidisciplinary introduction: Morgan & Claypool Publishers; 2008.

20. Shoham Y, Leyton-Brown K. Multiagent systems: Algorithmic, game-theoretic, and logical foundations: Cambridge University Press; 2008.

21. Efron B, Hastie T. Computer age statistical inference, student edition: algorithms, evidence, and data science: Cambridge University Press; 2021.

22. Marek K, Chowdhury S, Siderowf A, Lasch S, Coffey CS, Caspell-Garcia C, et al. The Parkinson’s progression markers initiative (PPMI)–establishing a PD biomarker cohort. Annals of clinical and translational neurology. 2018;5(12):1460–77.

23. Marek K, Jennings D, Lasch S, Siderowf A, Tanner C, Simuni T, et al. The Parkinson Progression Marker Initiative (PPMI). Prog Neurobiol. 2011;95(4):629–35.

24. Loo RTJ, Pavelka L, Mangone G, Khoury F, Vidailhet M, Corvol J-C, et al. Multi-cohort machine learning identifies predictors of cognitive impairment in Parkinson’s disease. npj Digital Medicine. 2025;8(1):482.

25. Almgren H, Camacho M, Hanganu A, Kibreab M, Camicioli R, Ismail Z, et al. Machine learning-based prediction of longitudinal cognitive decline in early Parkinson’s disease using multimodal features. Scientific reports. 2023;13(1):13193.

26. Simuni T, Merchant K, Brumm MC, Cho H, Caspell-Garcia C, Coffey CS, et al. Longitudinal clinical and biomarker characteristics of non-manifesting LRRK2 G2019S carriers in the PPMI cohort. npj Parkinson’s Disease. 2022;8(1):140.

27. U.S. Food and Drug Administration. Marketing Submission Recommendations for a Predetermined Change Control Plan for Artificial Intelligence-Enabled Device Software Functions: Guidance for Industry and Food and Drug Administration Staff. Guidance Document Silver Spring, MD: US Food and Drug Administration. 2025.

